# Trajectories and correlates of poor mental health in India over the course of the COVID-19 pandemic: a nation-wide survey

**DOI:** 10.1101/2023.09.13.23295513

**Authors:** Emma Nichols, Sarah Petrosyan, Pranali Khobragade, Joyita Banerjee, Marco Angrisani, Sharmistha Dey, Prof David E. Bloom, Simone Schaner, AB Dey, Jinkook Lee

## Abstract

**Introduction:** The COVID-19 pandemic had large impacts on mental health; however, most existing evidence is focused on the initial lockdown period and high-income contexts. By assessing trajectories of mental health symptoms in India over two years, we aim to understand the effect of later time periods and pandemic characteristics on mental health in a lower-middle income context.

**Methods:** We used data from the Real-Time Insights of COVID-19 in India (RTI COVID-India) cohort study (N=3,662). We used covariate-adjusted linear regression models with generalized estimating equations to assess associations between mental health (PHQ-4 score) and pandemic periods as well as pandemic characteristics (COVID-19 cases and deaths, government stringency, self-reported financial impact, COVID-19 infection in the household) and explored effect modification by age, gender, and rural/urban residence.

**Results:** Mental health symptoms dropped immediately following the lockdown period but rose again during the delta and omicron waves. Associations between mental health and later pandemic stages were stronger for adults 45 years of age and older (p<0.001). PHQ-4 scores were significantly and independently associated with all pandemic characteristics considered, including estimated COVID-19 deaths (PHQ-4 difference of 0.041 SD units; 95% Confidence Interval 0.030 - 0.053), government stringency index (0.060 SD units; 0.048 - 0.072), self-reported major financial impacts (0.45 SD units; 0.41-0.49), and COVID-19 infection in the household (0.11 SD units; 0.07-0.16).

**Conclusion:** While the lockdown period and associated financial stress had the largest mental health impacts on Indian adults, the effects of the pandemic on mental health persisted over time, especially among middle-age and older adults. Results highlight the importance of investments in mental health supports and services to address the consequences of cyclical waves of infections and disease burden due to COVID-19 or other emerging pandemics.

## INTRODUCTION

COVID-19 has had wide-reaching impacts since the WHO first declared COVID-19 as a global pandemic on March 11, 2020 [1]. In reaction to the initial spread of COVID-19, many countries implemented strict lockdown periods, closing businesses, schools, and requiring people to stay home [2,3]. Concerns over short- and long-term mental health consequences of the pandemic and lockdown period quickly emerged and were later backed up by quantitative evidence [4–8]. Although the majority of the empirical evidence comes from high-income settings, data from India and other low- and middle-income countries also highlight the impact of the initial lockdown period on mental health outcomes in these contexts [9,10].

Since the initial stages of the pandemic, the direct impacts of COVID-19 have continued to grow, with over 7.2 million reported and 17.7 million estimated deaths globally through Dec 1, 2022 [11]. The state of the pandemic has also evolved over time, with rapid shifts due to new variants, waves, and the development of vaccines. Despite potential impacts of the constantly changing landscape on mental health, longitudinal studies of mental health during the pandemic have been relatively rare.

Existing studies of mental health trajectories during the COVID-19 pandemic have predominantly focused on the initial lockdown period in 2020 and have been concentrated in high-income settings [12–16]. Evidence largely suggests that mental health symptoms peaked in the initial lockdown phase, and recovered as lockdown measures were lifted [12–16]. However, these studies are unable to make conclusions about the impacts of later waves and changes during the pandemic. Although fewer studies assessing later periods during the COVID-19 pandemic exist, some evidence from Argentina, Australia, Switzerland, and the US suggests that trends may be more complex, with both increases and decreases in mental health symptom burden during later periods of the pandemic [17–21].

Despite the large number of estimated COVID-19 infections and deaths in India [22,23], to our knowledge, only one prior study from India assessed mental health in the general population over more than two time periods [24]. Data showed that the initial lockdown period and the second wave (delta) led to similar levels of depression in rural but not urban settings [24]. However, the study was limited to two distinct regions in India and did not include follow-up beyond June 2021. As the impacts of COVID-19 continue to persist and evolve, it is important to understand the effects of later pandemic periods on mental health. The study of mental health outcomes throughout all stages of the pandemic in India also presents unique opportunities due to the presence of distinct pandemic phases coinciding with strict lockdown measures (initial phase), large numbers of pandemic-related deaths (delta wave), and high numbers of infections (omicron wave). The separation of these phases in time can enhance our understanding of the specific factors impacting mental health.

We aim to assess mental health over the course of the COVID-19 pandemic in India using data from the Real-Time Insights of COVID-19 in India (RTI COVID-India) survey, a nation-wide survey with 9 waves of data collection spanning from May 2020 to May 2022. By leveraging data over the course of the pandemic, we describe the impacts of various phases of the pandemic on mental health and identify characteristics of the pandemic most strongly associated with poor mental health. We also assess whether the impact of the pandemic and pandemic characteristics on mental health varies by age, gender, and urban/rural residence.

## METHODS

### Sample

The Real-Time Insights of COVID-19 in India (RTI COVID-India) survey included respondents aged 18-102 from 1,766 households across India [25]. The study sampled households from the Harmonized Diagnostic Assessment of Dementia for the Longitudinal Aging Study in India (LASI-DAD) study, a nationally representative sample of older adults 60 years and older from 18 states and union territories. Nine rounds of telephone-based surveys were administered from May 2020 to May 2022 (Figure S1). Respondents contributed an average of 5.6 (SD = 2.5) rounds of data collection, with data collected from a total of 21,108 phone surveys (N=3797). We excluded individuals without valid data on items from the Patient Health Questionnaire (PHQ-4) (N=666 surveys; N=83 respondents) or without valid data on pandemic characteristics of interest (N=1558 surveys; N=52 respondents). All participants provided informed verbal consent and data collection procedures were approved by Institutional Review Boards at the University of Southern California (study number UP-20-00277) and the All India Institute of Medical Sciences (study number RP-29/2020).

### Mental health measure

The PHQ-4 was used as a screening tool for anxiety and depression and was administered across all rounds [26]. The PHQ-4 asks respondents how often they have been bothered by: 1) feeling nervous, anxious, or on edge, 2) not being able to stop or control worrying, 3) feeling down, depressed, or hopeless, and 4) having little interest or pleasure in doing things. Due to the brief nature of the tool, we consider the four-item scale as a single marker of poor mental health. We estimated scores on this latent trait of poor mental health using a graded response item response theory model. These models relax the standard assumptions that all items contribute equally to total score and that differences between response options are equal. Variance in the factor represents the shared variance among the indicators and does not include random error specific to each item. Scores were scaled to have a mean of 0 and standard deviation of 1. Because respondents did not report any symptoms at a large proportion of surveys, we also estimated models for the presence of any mental health symptom as a binary outcome in sensitivity analyses.

### Pandemic measures

We assessed both reported and estimated state-level numbers of cases and deaths (per 100,000 population [from the 2011 Indian census]) due to COVID-19 in India. Reported administrative data from the Indian government was accessed at https://data.covid19bharat.org. Because official reported numbers are known to undercount numbers of cases and deaths, we also used estimated infections and excess mortality from the Institute of Health Metrics and Evaluation [11,22,23]. To capture the impact of lockdown measures, we used a state-specific index measure of government stringency, which captures the intensity of closings of public spaces and restrictions on gatherings and movement (details in Appendix 3) [27]. To facilitate comparisons between indicators, we converted them into z-scores. Because respondents’ mental health is likely affected by the conditions in the time period leading up to the survey, we created lagged indicators summarizing the mean level of cases, deaths, and government stringency over the time period leading to the interview date. We used a period of 3 months for the health-related indicators and a period of 1 month for the stringency index to reflect the fact that mental health impacts of health loss may last longer, while individuals may adapt to new policy environments more quickly. Based on the real-time and lagged metrics of cases, deaths, and government response, the Indian Council of Medical Research COVID-19 timeline, and limited data on variant seroprevalence trends, we divided the COVID-19 pandemic into 6 time periods: lockdown, first wave, low cases (winter 2020/21), delta wave, low cases (fall 2021), and omicron wave (Figure 1) [28,29].

**Figure 1.**
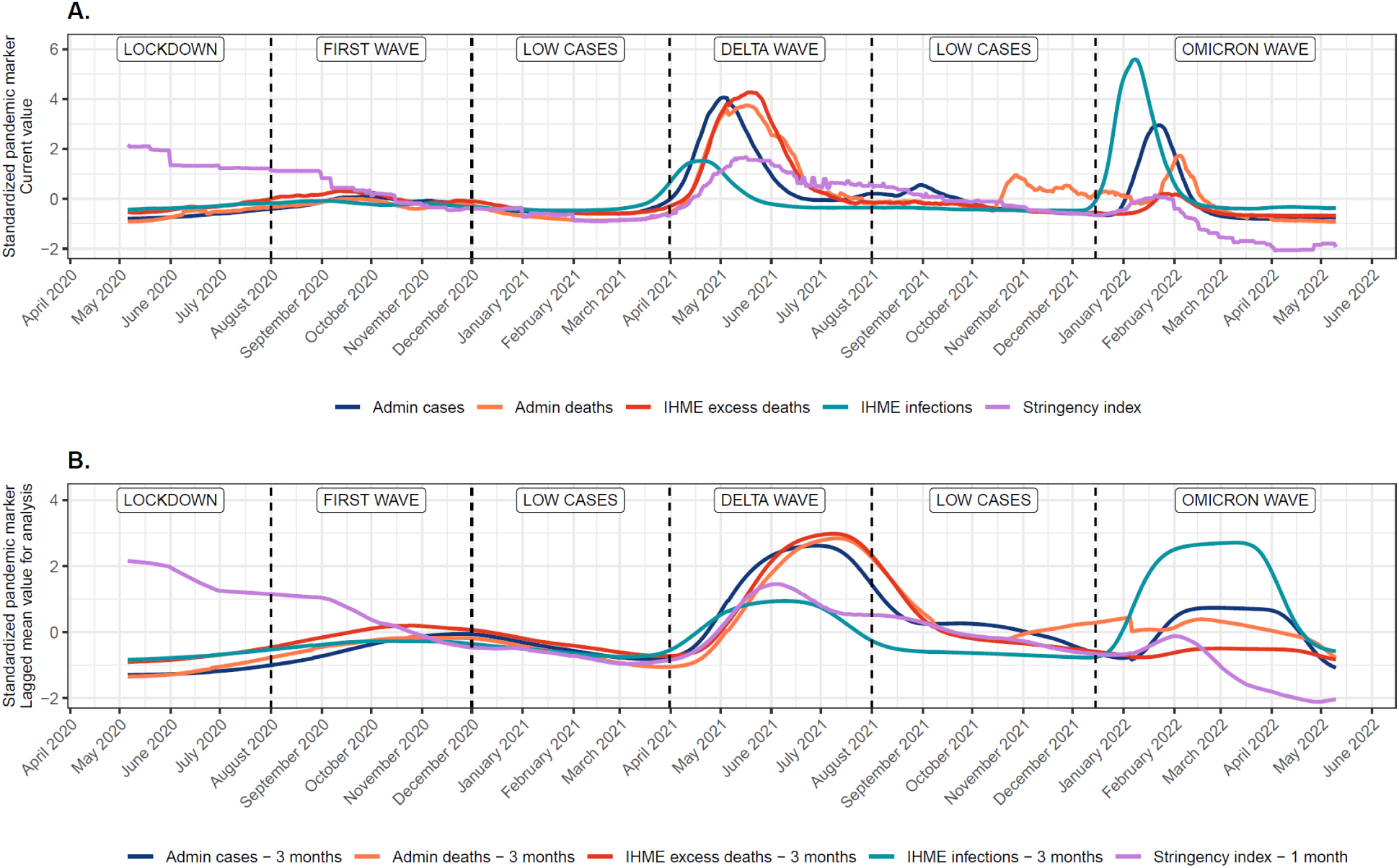
Trajectories in pandemic markers over the course of the COVID-19 pandemic during the time frame of data collection for the Real-Time Insights of COVID-19 in India study. All pandemic markers are standardized to z-scores to facilitate comparisons in trajectories over time. All metrics are reported or estimated at the state level; overall levels shown here are weighted based on the distribution of participants across the Indian states and territories. Panel A shows current values of pandemic markers; Panel B shows mean pandemic markers over the time periods used for analysis (3 months of case and death indicators, 1 month for government stringency index).

We used prior household infection as an indicator of the direct impact and experience of COVID-19 infection. In constructing this indicator, we considered information from reports of diagnosed infections from any household member in waves 2-8, reports of confirmed or suspected infections from any household member in waves 7-8, and dates of reported or suspected household infections from a single female household member in wave 9. Finally, we used data on self-reported financial impact of the pandemic (no impact/minor impact/moderate impact/major impact/too soon to tell) reported by a male household member to assess the effect of perceived financial distress.

### Covariates

We considered self-reported age, gender, and educational attainment (none/less than secondary/secondary/some graduate) from survey data as well as rural/urban residence as covariates.

### Statistical analysis

We used descriptive statistics (proportions and numbers for binary variables, means and interquartile ranges for continuous variables) to characterize included covariates and pandemic markers over the 6 pandemic periods and 9 waves considered in analyses. We also visualized trajectories of PHQ-4 score over time. To assess the association between each pandemic indicator and mental health, we used linear regression models estimated with generalized estimated equations (GEE) with an exchangeable correlation structure to account for correlation attributable to the inclusion of multiple observations for a given respondent. For each indicator, we estimated unadjusted models and models adjusted for age (estimated with a natural cubic spline with 2 internal knots [33^rd^ and 66^th^ percentiles] and 2 external knots [5^th^ and 95^th^ percentile]), gender, educational attainment, and rural/urban residence. To understand individual contributions of each pandemic indicator while adjusting for other indicators, we estimated a single GEE model including covariates as well as all pandemic characteristics except time period and administrative cases and deaths. We chose to use estimated rather than reported cases and deaths in this single model to avoid bias due to under-reporting particularly in later stages of the pandemic.

Finally, we assessed effect modification of the associations between each pandemic characteristic and mental health by age category (<30/30-44/45-59/60-74/75+), gender, and rural/urban residence.

Stratified results to visualize effect modification were estimated using linear combinations of coefficients from GEE models including an interaction between the pandemic characteristic and effect modifiers of interest. Models controlled for the same covariates used in previous models but excluded any covariate considered as an effect modifier. Chi-square tests comparing models with an interaction to models without an interaction were used to test the overall statistical significance of effect modification. In sensitivity analyses, we re-estimated all models described above using a binary indicator for the presence of any mental health symptom. For binary models, we used Poisson regression with robust variance to estimate prevalence ratios.

All descriptive statistics used survey weights to correct for sampling processes and selection bias; we present unweighted regression models in main analyses and weighted regression models in sensitivity analyses. Item response theory models were estimated in Mplus Version 8; all other models were estimated in R version 4.2.2.

## RESULTS

We included data from 3,662 respondents collected across 18,884 phone surveys. Demographics, including age, gender, educational attainment, and rural/urban residence were stable across wave and pandemic period (Table 1, Table S1-2). Although reported cases were similar between the delta and omicron waves, estimated cases were higher during the omicron wave. Reported and estimated deaths were higher during the delta wave. Government stringency index was highest during the lockdown phase, while self-reported financial impact did not vary substantially over time. PHQ-4 score varied over time and was highest during the lockdown phase and at the end of the delta wave (Table 1, Figure 2). All age groups had similarly high PHQ-4 scores in the initial phases of the pandemic but in later stages of the pandemic, PHQ-4 sores were higher among the older age groups (Figure 2).

**Table 1.**
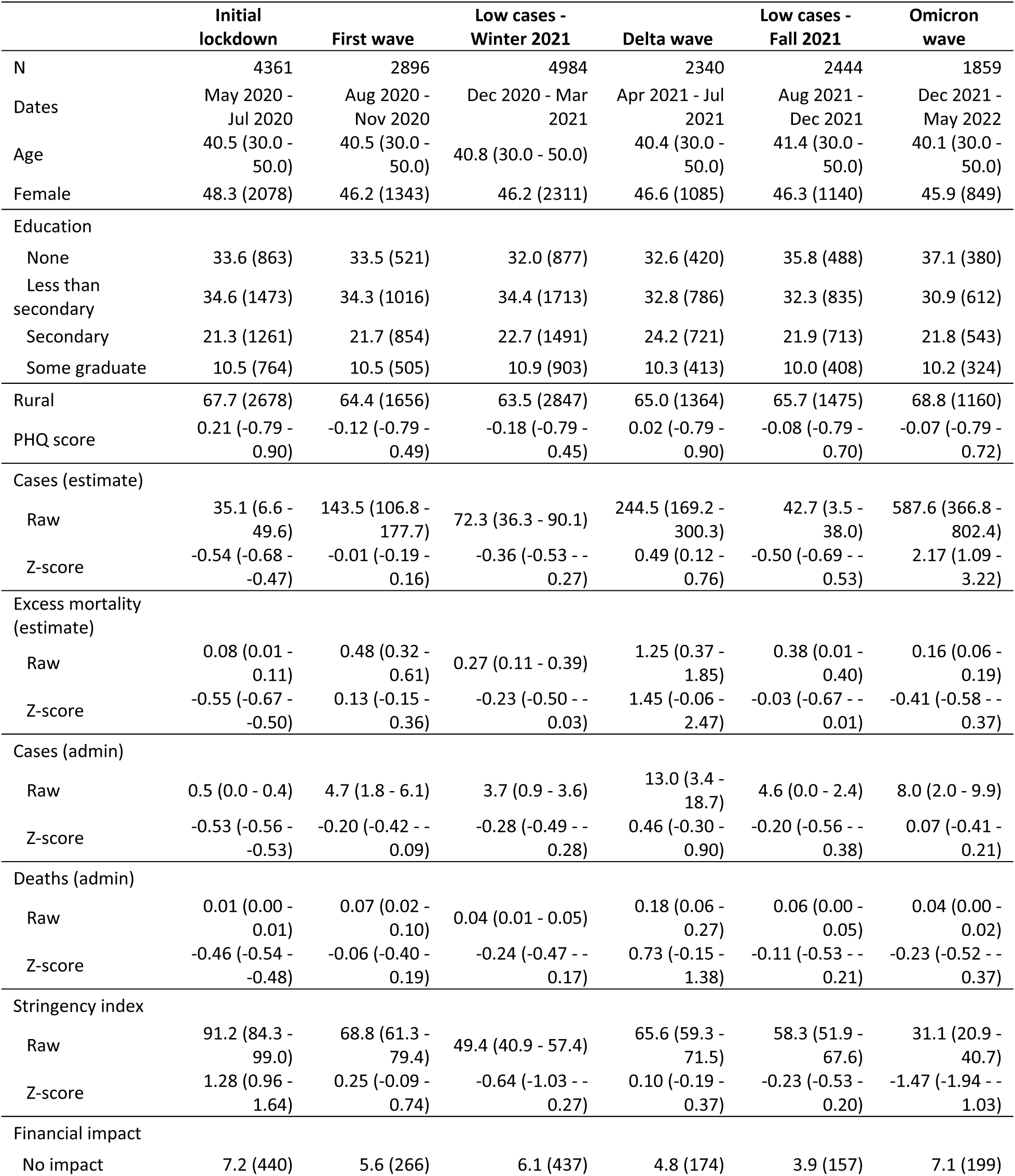
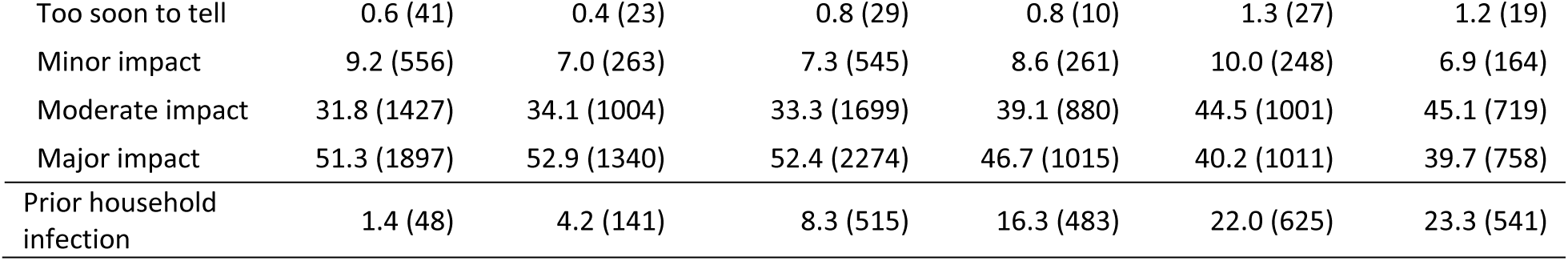
Characteristics of the pandemic and the Real-Time Insights of COVID-19 in India sample (N=3,662) across the 6 pandemic time periods. Binary and categorical variables are presented using weighted proportions and numbers of respondents; continuous variables are presented using weighted means and interquartile ranges. Cases, deaths, and stringency index shown summarize the mean lagged values (over the three [cases, deaths] months or one [government stringency] month prior to interview date) for all those interviewed during a given pandemic period. Raw cases and deaths are presented per 100,000 population.

**Figure 2.**
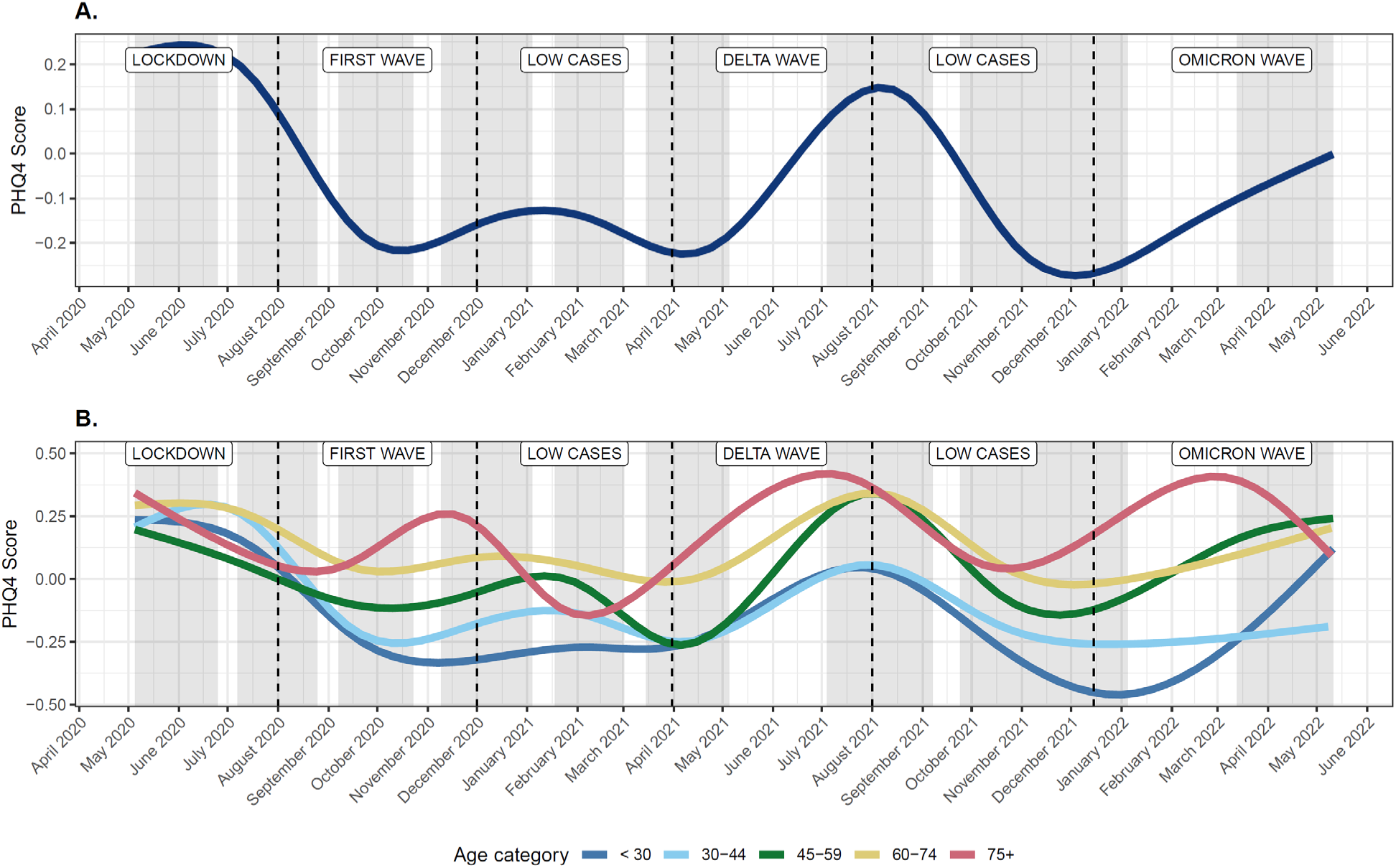
Estimated smoothed trajectories of PHQ4 scores for participants in the Real-Time Insights of COVID-19 in India sample (N=3,662) over the course of the pandemic, overall (A) and by age category (B). Trajectories were fit with generalized additive models with flexible penalized cubic splines. Grey shaded regions reflect time periods with ongoing data collection.

Crude and adjusted associations between pandemic periods from regression models were similar and suggested that mean PHQ-4 scores in all subsequent pandemic periods were lower than mean PHQ-4 score during the lockdown phase (Figure 3). However, there was variation over time, and in adjusted models mean PHQ-4 score during the delta wave was only slightly lower than during the lockdown period (difference of -0.06 95% Confidence Interval [CI] -0.10- -0.02 SD units). PHQ-4 scores were also higher during the omicron wave (difference of -0.10; -0.15- -0.05 SD units compared to lockdown) as compared to the period immediately following the initial lockdown period (first wave [-0.22; -0.26- - 0.19] and Winter 2021 [-0.23; -0.26- -0.20]).

**Figure 3.**
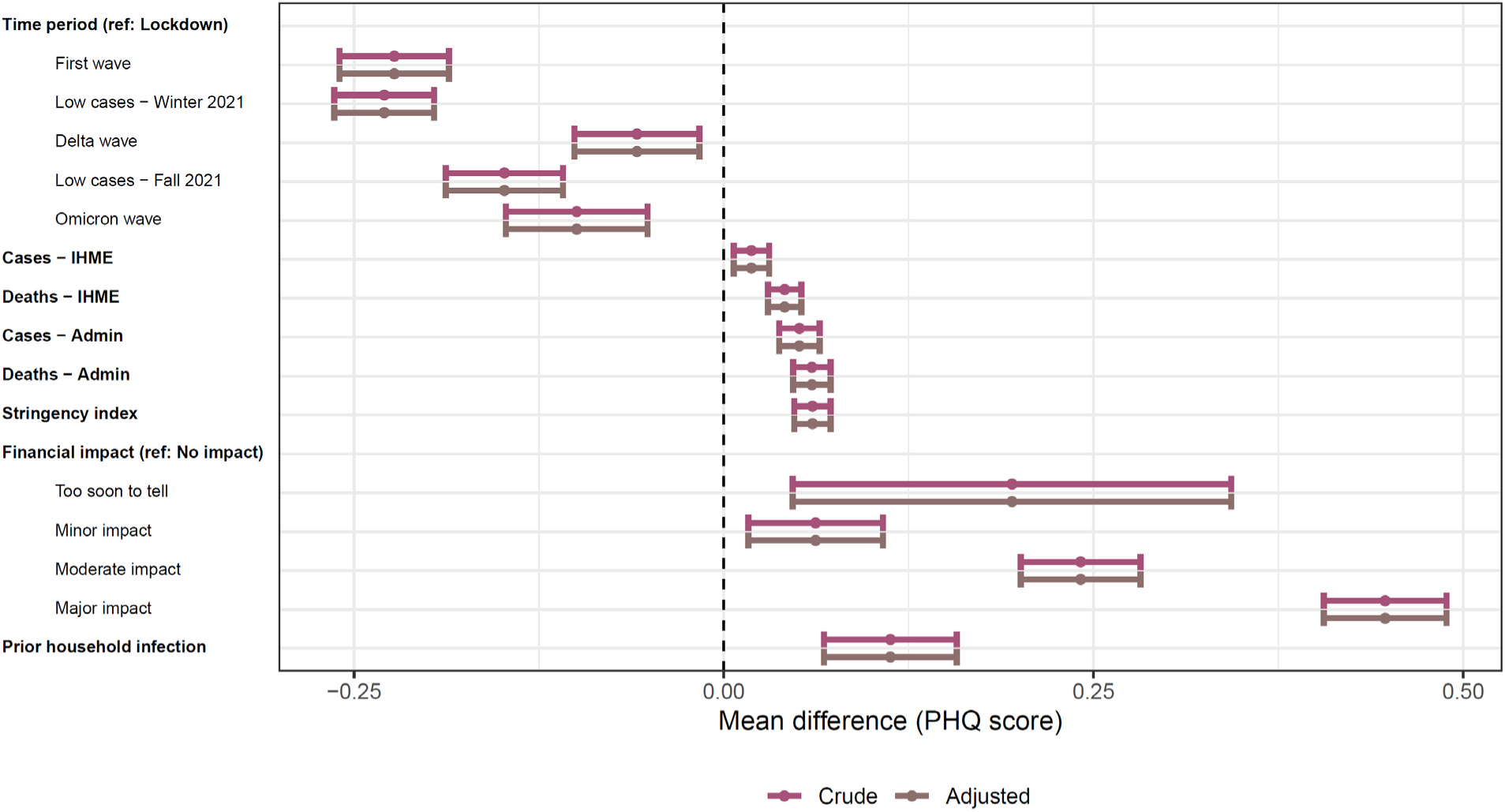
Associations between COVID-19 pandemic periods and COVID-19 pandemic markers and PHQ-4 scores in the Real-Time Insights of COVID-19 in India sample (N=3,662). Associations with all continuous markers (cases, deaths, government stringency index) are shown as associations for 1 SD unit difference. Adjusted models are adjusted for age (spline), gender, educational attainment, and rural/urban residence.

Estimated case counts were positively associated with PHQ-4 score, such that a 1 SD higher estimated case count was associated with 0.02 (95% CI 0.01-0.03) SD higher PHQ-4 score (Figure 3). The estimated association between reported case counts and PHQ-4 score was larger (0.05 [0.04-0.06]), likely because cases were more likely to be underreported during the later stages of the pandemic when the disease was less severe. Reported and estimated deaths as well as government stringency index were positively associated with PHQ-4 score; mean effect sizes ranged from 0.04 to 0.06 SD units. The estimated association between mental health and prior household infection (0.11; 0.07-0.16) was larger than the estimated association between mental health and contextual pandemic markers; however, the difference between the magnitude of coefficients was not statistically significant as the estimated association for prior household infection was imprecise. The largest estimated association was observed for self-reported financial impact; compared to those reporting no impact, those reporting a major financial impact had 0.45 (0.41-0.49) SD higher PHQ-4 scores. Individual associations for pandemic markers remained stable when estimated in a single model (Table S3), indicating that associations between each pandemic marker and PHQ-4 score were largely independent.

Age was an important effect modifier of the association between pandemic period and pandemic markers and PHQ-4 scores (Figure 4); adding interaction terms significantly improved model fit for all pandemic markers (p<0.001) (Table S4). PHQ-4 scores were more evenly elevated over the course of the entire pandemic in respondents 45 years of age and older, whereas in younger respondents (under 44 years of age), PHQ-4 scores dropped and stayed lower after the initial lockdown period (Figure 4, Table S4). Associations between PHQ-4 scores and pandemic markers including self-reported financial impact, cases, deaths, and prior household infection were stronger among older respondents (for all: p<0.001). However, the opposite was true for government stringency index. Gender differences were largest for pandemic period (p=0.003), and financial impact (p<0.001); there were stronger associations between the lockdown period and financial effects of the pandemic for men compared to women. Additionally, PHQ-4 score was more strongly associated with later pandemic periods (omicron and delta waves) (p=0.037), and the association with estimated deaths was stronger (p<0.001) in urban compared to rural areas (Figure 4, Table S4).

**Figure 4.**
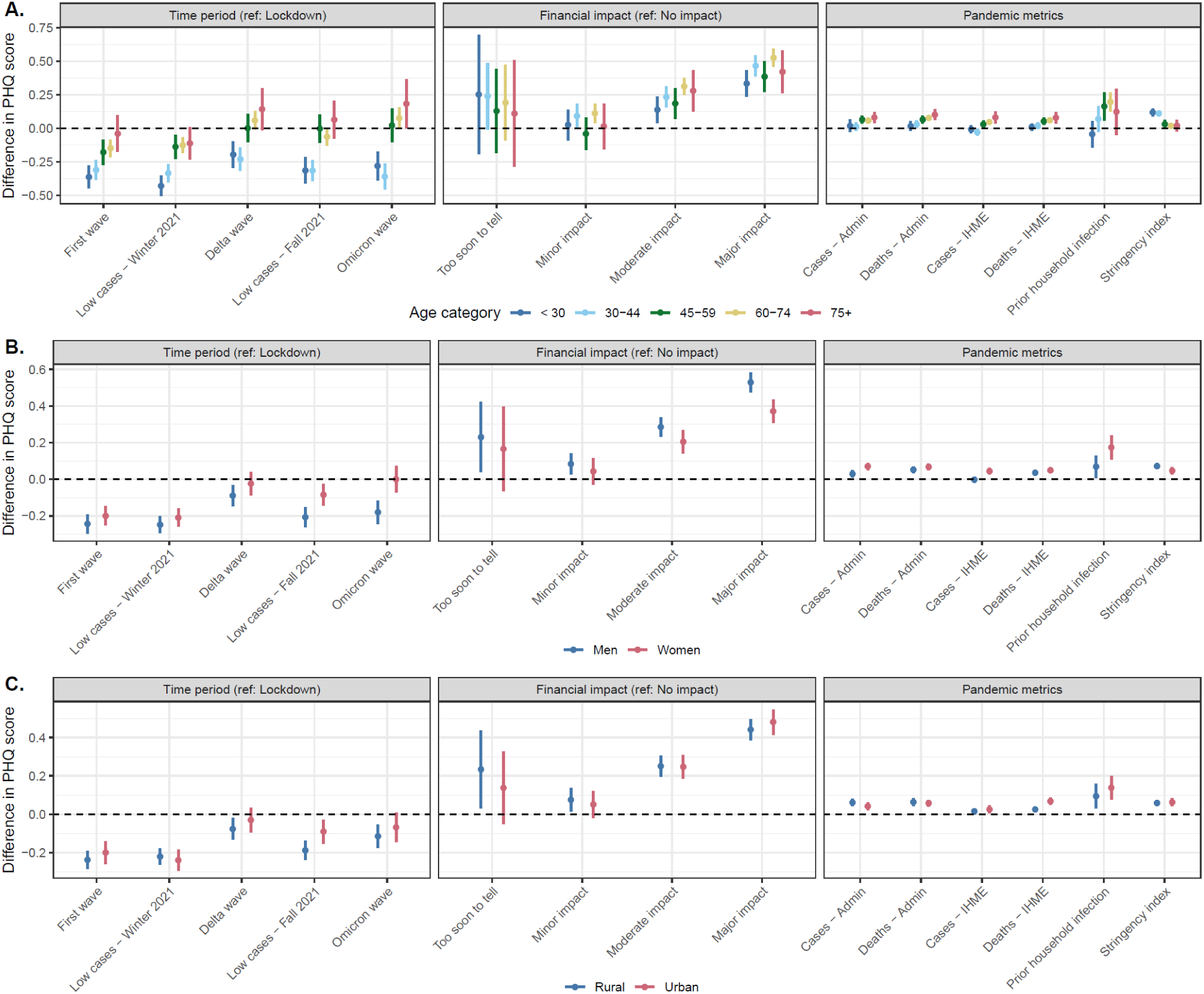
Effect modification of the association between COVID-19 pandemic periods and COVID-19 pandemic markers and PHQ-4 scores by age group in the Real-Time Insights of COVID-19 in India sample (N=3,662). All models are adjusted for gender, educational attainment, and rural/urban residence.

Results from models using the binary outcome of any mental health symptoms followed similar patterns (Figures S2-3, Table S5). Results from weighted regression models were closer to results reported for younger participants, as the RTI COVID-India study oversampled older adults compared to the general population of India (Figures S4-5, Table S6).

## DISCUSSION

In a nation-wide sample from India, mental health symptoms fluctuated over the course of the pandemic. High mental health symptom burden was especially apparent during the lockdown period, but symptom burden was also high at later points in the pandemic in response to COVID-19 waves (omicron, delta), particularly among older respondents (aged 45 and older). Estimated and reported cases, estimated and reported deaths, government stringency, self-reported financial burden, and reported infection in the household were all associated with poorer mental health.

Our results on mental health during the lockdown period align with a large number of studies that have reported high mental health symptom burden in the initial lockdown stage of the pandemic in India [30,31]. The literature on mental health outcomes in later stages of the pandemic in India is more scarce and somewhat mixed; studies have reported both increasing and decreasing symptom burden over time [24,32,33]. Our findings suggest that such inconsistent evidence could be due to differences in the timing of survey waves and longitudinal follow-ups; we found both increasing and decreasing symptom burden during different time periods. The number of assessments and length of follow-up time allowed us to link subsequent waves of COVID-19 and impacts of COVID-19 (e.g., deaths) to mental health outcomes. In other contexts, including Australia, Poland, the US, and the United Kingdom, prior studies have also found evidence of poorer mental health outcomes in response to later COVID-19 waves, in line with our findings [20,34–36].

We were also able to assess correlates of poor mental health and compare the relative magnitude of associations across different pandemic attributes. Significant associations across all attributes considered suggests that such characteristics could help policy-makers target time-periods or regions with greater need for mental health supports and services. Similarly to other studies across contexts, we found that financial stress was strongly associated with poor mental health [37–39]. In our study, perceived pandemic-induced financial burden had the strongest associations with poor mental health of the pandemic characteristics considered. This finding aligns with prior work suggesting that perceived financial burden is an important mediator of the effects of job and income loss on mental health [40].

Although policies affecting financial stress were largely concentrated in the early lockdown period, our data indicated that perceived financial stress was high over the course of the COVID-19 pandemic in India. Financial stress may have larger consequences in settings such as India due to higher levels of poverty and food insecurity compared to other settings [41].

Although the financial effects of government lockdown policies have been previously emphasized [42], we found that the association between government stringency index and poor mental health outcomes was independent of perceived financial stress. The independence of the effects of government stringency and financial distress suggests that social and psychosocial factors impacted by lockdown, including loneliness and fear, may also play an important role [9,43]. Prior cross-national research has also found an association between policy stringency and poor mental health [44]. However, other work suggests that prompt implementation of restrictions with clear messaging may actually be beneficial [45]. Considered together, this evidence stresses the importance of effective and quick communication and action. In contrast, delayed or draw-out lockdown periods with poor communication, as was experienced in India [46], are more likely to have negative impacts on mental health. Additionally, in our study associations between policy stringency and mental health were strongest for younger participants and were attenuated in older adults, who may feel more protected in more controlled environments.

We also found that increases in reported and estimated cases and deaths, and COVID-19 infection in the household were associated with poorer mental health. Prior studies in the US have also documented associations between reported cases and worse mental health outcomes, however, these studies did not compare associations for reported versus estimated infections [47,48]. In our study, the association between COVID-19 deaths and mental health was stronger in urban areas, where mass open-air funeral pyres and higher population density may have magnified fear and awareness of potential negative consequences of COVID-19 [49]. Overall, results indicate that the presence of severe disease likely has negative effects on mental health, which should be considered when quantifying and summarising the overall impact of COVID-19.

Differences in trajectories of mental health and associations between pandemic characteristics and mental health across age groups point to differences in the experience of the pandemic across age. While mental health symptoms were elevated across all age groups during the initial lockdown phase, during later stages of the pandemic, elevated symptoms were observed primarily among adults 45 years and older. Furthermore, associations between measures of pandemic burden (cases, deaths) were also stronger among those 45 years and older, while they were largely null among younger adults. Prior evidence comparing the pre-pandemic to lockdown periods indicated that effects of the pandemic on mental health were larger for younger adults [14,16]. However, based on this study, conclusions that older adults were able to cope with the COVID-19 pandemic better than younger adults may have been premature [50]. Instead, current evidence from India suggests that the mental health impacts of pandemic burden (household infections, deaths) after the initial lockdown phase may be larger among older adults.

This study is the first large-scale nation-wide effort to assess mental health throughout the COVID-19 pandemic in India. Because of the design, scale, and setting of the study, we were able to disentangle associations between a variety of different pandemic characteristics and poor mental health. In the context of study strengths, limitations should be considered. First, the implementation of telephone assessments may have introduced selection bias. Although India has high rates of mobile phone ownership (93% of households), there are gaps by gender and factors related to socio-economic status [51]. To adjust for this potential bias, we used survey weights adjusted for non-response in sensitivity analyses; weight construction included a post-stratification raking algorithm to ensure the weighted sample matched national population demographics. Second, due to concerns about participant burden we were unable to implement detailed measures of mental health across all time points. Instead, we used the PHQ-4, which is commonly used as a screening tool for anxiety and depression. The scale only includes four items and a large proportion of the sample reported zero symptoms. However, we used item-response theory methods to estimate latent mental health with maximal precision, and patterns of results were consistent in models assessing trajectories and correlates of reporting any mental health symptom. Despite its brevity, the PHQ-4 correlates strongly with more comprehensive measures of mental health and shows strong criterion and construct validity [26]. Third, participant dropout between study waves may bias estimates. However, we would hypothesize that those who drop out would be more likely to have worse mental health outcomes in response to pandemic stressors; in this scenario, our results including only those retained in the study are conservative. Additionally, analysis of reasons for drop-out indicates that over 60% of households who dropped out of the study for a given wave dropped out due to reasons that are less likely to be due to mental health (e.g., phone call not answered, phone number changed or not in network) compared to refusal or withdrawal of consent.

Finally, we were unable to examine the impacts of the vaccination campaign on mental health because of the strong correlation in time between the delta wave and vaccination; other contexts may be better suited to tease out these associations.

Overall, this study highlighted the complexity of mental health trajectories over the course of the COVID-19 pandemic in India. The initial lockdown period was associated with a high burden of mental health symptoms, but subsequent pandemic waves and associated mortality or household infections were also associated with worsening mental health, particularly among middle-aged and older adults. Results suggest that the quantification of the consequences of COVID-19 throughout the pandemic should include the mental health consequences of COVID-19 in both the early and later stages of the pandemic. Our findings further highlight the importance of continued attention to mental health in the context of COVID-19 and future emerging pandemics and support the need for ongoing services and interventions aimed at supporting mental health in the general population.

## Data Availability

Data are available to download via the Gateway to Global Aging website: covid.g2aging.org. Users must sign a data use agreement before being granted access to the data.

https://www.covid.g2aging.org

## Acknowledgements: We acknowledge all members of the study team who made data collection possible. We also acknowledge the contribution of all participants who consented to take part in this study during the COVID-19 pandemic.

Declaration of interests: None.

Funding: National Institute on Aging, National Institutes of Health (U01AG065958, R01AG051125, R01 AG030153).

Data availability statement: Data are available to download via the Gateway to Global Aging website: covid.g2aging.org. Users must sign a data use agreement before being granted access to the data.

