## Appendix for "Trajectories and correlates of poor mental health in India over the course of the COVID-19 pandemic: a nation-wide survey"

#### Appendix 1: Timing of participant responses by wave over the COVID-19 pandemic

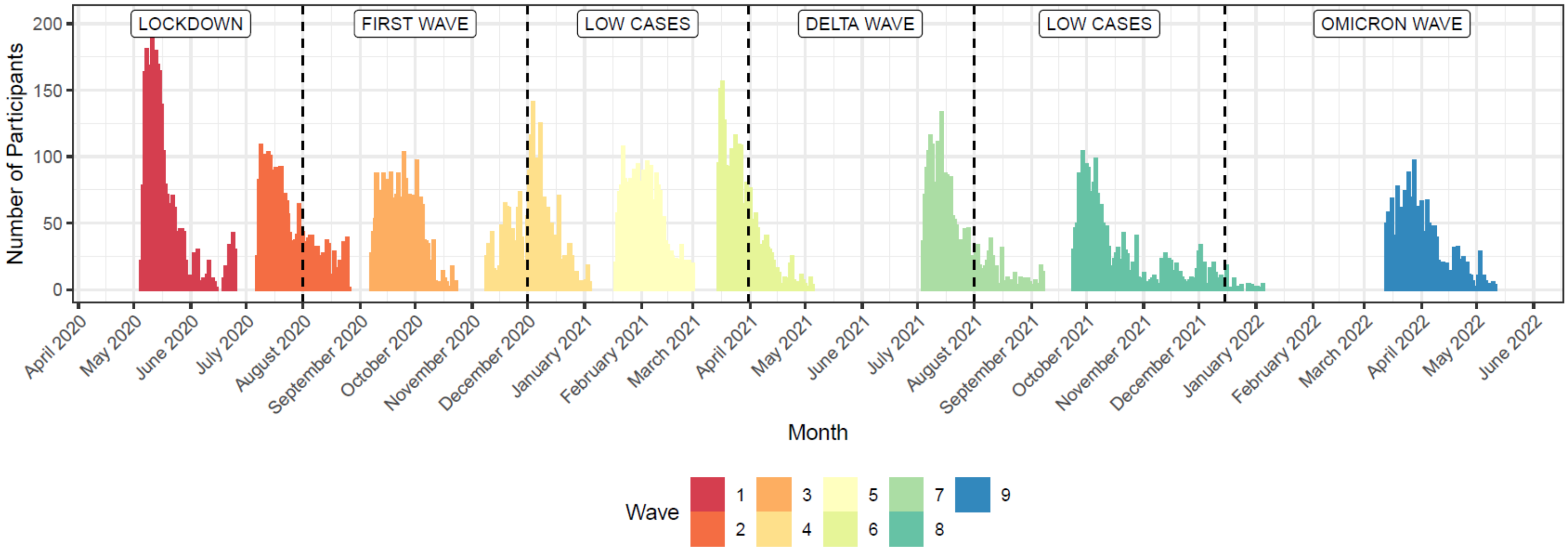

**Figure S1. Timing of participant responses by wave over the COVID-19 pandemic**

#### Appendix 2: Construction of the Government Stringency Index

The government stringency index was developed by the Oxford COVID-19 Government Response tracker (OxCGRT) team. This below information and additional details are available in the summary paper (Hale T, Angrist N, Goldszmidt R, et al. A global panel database of pandemic policies (Oxford COVID-19 Government Response Tracker). Nat Hum Behav 2021; 5: 529–38.) and at the GitHub data repository (https://github.com/OxCGRT/covid-policy-tracker/). The stringency index summarizes data across 9 indicators, listed below along with the possible response options:

- School and university closings: 0 = no measures, 1 = recommended closing or all schools open with alternations resulting in significant differences compared to non-COVID-19 operations, 2 = require closing (only some levels or categories such as just high school or just public schools), 3 = require closing all levels
- Workplace closings: 0 = no measures, 1 = recommended closing (or work from home) or all businesses open with alternations resulting in significant differences compared to non-COVID-19 operations, 2 = require closing (or work from home) for some sectors or categories of workers, 3 = require closing (or work from home) for all-but essential workplaces (grocery stores, doctors)
- Cancel public events: 0 = no measures, 1 = recommend cancelling, 2 = require cancelling
- Limits on gatherings: 0 = no restrictions, 1 = restrictions on very large gatherings (limit above 1000), 2 = restrictions on gatherings between 101-1000 people, 3 = restrictions on gatherings between 11-100 people, 4 = restrictions on gathers of 10 people or less
- Closing public transport: 0 = no measures, 1 = recommend closing (or significantly reduce volume/route/means of transport available), 3 = require closing (or prohibit most citizens from using it)
- Orders to “shelter-in place” and otherwise confine to the home: 0 = no measures, 1 = recommend not leaving house, 2 = require not leaving house with exceptions for daily exercise, grocery shopping, and ‘essential’ trips, 3 = require not leaving house with minimal exceptions (e.g., allowed to leave once a week or only one person can leave at a time)
- Restrictions on internal movement between cities/regions: 0 = no measures, 1 = recommend not to travel between regions/cities, 2 = internal movement restrictions in place
- Restrictions on international travel: 0 = no restrictions, 1 = screening arrivals, 2 = quarantine arrivals from some or all regions, 3 = ban arrivals from some regions, 4 = ban on all regions or total border closure
- Presence of public info campaigns: 0 = no COVID-19 public information campaign, 1 = Public officials urging caution about COVID-19, 2 = coordinated public information campaign (e.g., across traditional and social media)

Each indicator also has a corresponding flag variable, indicating whether the policy applied to the entire state, or to a specific and constrained geographic area within that state. Indices were calculated by taking the average of the scaled score on each indicator, calculated as the score/maximal score*100. When a specific indicator was flagged for being geographically constrained, 0.5 was subtracted from the score before scaling. Where data for individual indicators was missing, the indicator was assumed to be 0, leading to a more conservative estimate. For the time period after vaccination was introduced, when policies differed by vaccination status, the stringency index was computed for both the vaccinated and unvaccinated populations, and the overall stringency index was estimated as the average of the vaccinated and unvaccinated indices.

#### Appendix 3: Characteristics of the sample and the pandemic across the 9 waves of data collection

**Table S1.** Characteristics of the pandemic and the Real-Time Insights COVID-19 in India sample (N=3,662) across the 9 waves of data collection. Binary and categorical variables are presented using weighted proportions and numbers of participants; continuous variables are presented using weighted means and interquartile ranges. Cases, deaths, and stringency index shown summarize the mean lagged values (over the three [cases, deaths] months or one [government stringency] month prior to interview date) for all those interviewed during a given pandemic period. Raw cases and deaths are presented per 100,000 population.

| | **Wave 1** | **Wave 2** | **Wave 3** | **Wave 4** | **Wave 5** | **Wave 6** | **Wave 7** | **Wave 8** | **Wave 9** |
|---|---|---|---|---|---|---|---|---|---|
| N | 2754 | 2193 | 1579 | 2041 | 2164 | 2154 | 2128 | 2061 | 1810 |
| Dates | May 2020 - Jun 2020 | Jul 2020 - Aug 2020 | Sep 2020 - Oct 2020 | Nov 2020 - Jan 2021 | Jan 2021 - Mar 2021 | Mar 2021 - May 2021 | Jul 2021 - Sep 2021 | Sep 2021 - Jan 2022 | Mar 2022 - May 2022 |
| Age | 40.6 (30.0 - 50.0) | 40.7 (30.0 - 50.0) | 40.0 (30.0 - 48.0) | 40.9 (30.0 - 50.0) | 40.7 (30.0 - 49.0) | 41.1 (30.0 - 50.0) | 40.4 (30.0 - 49.0) | 41.1 (30.0 - 50.0) | 40.1 (30.0 - 50.0) |
| Female | 49.5 (1334) | 47.1 (1039) | 44.3 (707) | 46.1 (927) | 46.3 (1002) | 46.5 (1008) | 47.1 (992) | 46.5 (969) | 45.8 (828) |
| Education | | | | | | | | | |
| None | 34.0 (542) | 35.0 (440) | 31.1 (283) | 32.1 (362) | 32.3 (389) | 33.2 (392) | 34.1 (383) | 34.3 (391) | 37.0 (367) |
| Less than secondary | 34.6 (938) | 33.5 (747) | 36.0 (545) | 33.3 (690) | 34.7 (755) | 32.9 (737) | 33.7 (731) | 32.3 (693) | 31.1 (599) |
| Secondary | 20.7 (786) | 21.5 (639) | 22.0 (458) | 23.3 (617) | 22.6 (640) | 23.5 (653) | 22.1 (637) | 22.9 (622) | 21.9 (531) |
| Some graduate | 10.7 (488) | 10.0 (367) | 10.9 (293) | 11.2 (372) | 10.4 (380) | 10.4 (372) | 10.2 (377) | 10.4 (355) | 10.0 (313) |
| Rural | 64.9 (1626) | 67.7 (1337) | 66.4 (932) | 64.5 (1179) | 65.5 (1281) | 63.7 (1230) | 65.2 (1265) | 64.2 (1202) | 69.0 (1128) |
| PHQ score | 0.22 (-0.79 - 0.90) | 0.15 (-0.79 - 0.72) | -0.22 (-0.79 - 0.32) | -0.15 (-0.79 - 0.49) | -0.12 (-0.79 - 0.52) | -0.22 (-0.79 - 0.32) | 0.11 (-0.79 - 0.90) | -0.13 (-0.79 - 0.52) | -0.06 (-0.79 - 0.72) |
| Cases (estimate) | | | | | | | | | |
| Raw | 14.6 (3.6 - 18.3) | 75.5 (47.8 - 84.9) | 156.5 (140.4 - 181.0) | 130.5 (78.6 - 158.6) | 57.9 (34.1 - 76.6) | 92.1 (32.6 - 123.3) | 229.5 (142.5 - 300.3) | 29.6 (2.8 - 30.0) | 603.6 (434.5 - 804.6) |
| Z-score | -0.64 (-0.69 - - 0.62) | -0.34 (-0.48 - -0.30) | 0.06 (-0.02 - 0.18) | -0.07 (-0.33 - 0.07) | -0.43 (-0.54 - -0.34) | -0.26 (-0.55 - - 0.11) | 0.41 (-0.01 - 0.76) | -0.57 (-0.70 - -0.56) | 2.25 (1.42 - 3.23) |
| Excess mortality (estimate) | | | | | | | | | |
| Raw | 0.03 (0.00 - 0.03) | 0.21 (0.10 - 0.30) | 0.51 (0.33 - 0.73) | 0.48 (0.27 - 0.66) | 0.25 (0.14 - 0.35) | 0.13 (0.06 - 0.13) | 1.60 (1.09 - 1.91) | 0.16 (0.01 - 0.16) | 0.17 (0.06 - 0.19) |
| Z-score | -0.64 (-0.68 - - 0.64) | -0.34 (-0.52 - -0.18) | 0.19 (-0.12 - 0.56) | 0.14 (-0.23 - 0.45) | -0.25 (-0.46 - -0.09) | -0.47 (-0.59 - - 0.46) | 2.04 (1.18 - 2.56) | -0.42 (-0.68 - -0.41) | -0.40 (-0.58 - - 0.36) |
| Cases (admin) | | | | | | | | | |

| | | | | | | | | | |
|---|---|---|---|---|---|---|---|---|---|
| Raw | 0.1 (0.0 - 0.1) | 1.5 (0.3 - 1.6) | 4.9 (1.9 - 7.7) | 6.1 (2.1 - 7.0) | 3.1 (1.2 - 2.8) | 2.5 (0.6 - 2.1) | 14.8 (5.4 - 20.4) | 3.7 (0.0 - 1.3) | 8.2 (2.0 - 9.9) |
| Z-score | -0.56 (-0.56 - -0.56) | -0.45 (-0.54 - -0.44) | -0.18 (-0.42 - 0.04) | -0.08 (-0.40 - -0.01) | -0.32 (-0.47 - -0.34) | -0.37 (-0.52 - -0.40) | 0.60 (-0.14 - 1.04) | -0.27 (-0.56 - -0.46) | 0.08 (-0.41 - 0.21) |
| Deaths (admin) | | | | | | | | | |
| Raw | 0.00 (0.00 - 0.00) | 0.03 (0.01 - 0.04) | 0.07 (0.02 - 0.13) | 0.07 (0.02 - 0.09) | 0.04 (0.01 - 0.05) | 0.02 (0.00 - 0.03) | 0.23 (0.08 - 0.32) | 0.03 (0.00 - 0.03) | 0.05 (0.00 - 0.02) |
| Z-score | -0.53 (-0.54 - -0.53) | -0.31 (-0.49 - -0.28) | -0.03 (-0.38 - 0.34) | -0.02 (-0.40 - 0.10) | -0.26 (-0.47 - -0.19) | -0.40 (-0.51 - -0.35) | 1.06 (0.05 - 1.68) | -0.30 (-0.54 - -0.36) | -0.23 (-0.52 - -0.37) |
| Stringency index | | | | | | | | | |
| Raw | 97.0 (96.1 - 99.4) | 81.5 (77.3 - 84.9) | 70.9 (63.7 - 78.4) | 53.8 (48.3 - 63.0) | 48.6 (40.8 - 54.2) | 48.8 (41.7 - 59.3) | 68.7 (64.6 - 74.3) | 56.5 (50.1 - 66.4) | 30.6 (20.7 - 40.1) |
| Z-score | 1.55 (1.50 - 1.66) | 0.84 (0.64 - 0.99) | 0.35 (0.02 - 0.69) | -0.43 (-0.69 - -0.02) | -0.67 (-1.03 - -0.42) | -0.67 (-0.99 - -0.19) | 0.25 (0.06 - 0.51) | -0.31 (-0.61 - 0.14) | -1.50 (-1.95 - -1.06) |
| Financial impact | | | | | | | | | |
| No impact | 8.2 (301) | 4.8 (187) | 6.5 (157) | 4.2 (161) | 7.4 (226) | 5.4 (153) | 4.4 (142) | 4.6 (147) | 7.3 (199) |
| Too soon to tell | 0.6 (20) | 0.4 (26) | 0.7 (15) | 0.4 (11) | 0.6 (5) | 0.9 (16) | 1.9 (27) | 0.6 (10) | 1.3 (19) |
| Minor impact | 8.8 (366) | 9.6 (253) | 6.9 (141) | 6.7 (173) | 7.5 (262) | 9.2 (272) | 8.1 (219) | 9.3 (192) | 6.7 (159) |
| Moderate impact | 31.2 (885) | 34.0 (769) | 32.5 (549) | 30.7 (607) | 30.8 (673) | 39.9 (884) | 41.3 (830) | 44.6 (840) | 45.1 (693) |
| Major impact | 51.2 (1182) | 51.1 (958) | 53.4 (717) | 58.0 (1089) | 53.6 (998) | 44.5 (829) | 44.3 (910) | 41.0 (872) | 39.7 (740) |
| Prior household infection | 0.6 (9) | 2.8 (55) | 3.6 (62) | 6.7 (168) | 7.6 (211) | 9.8 (259) | 19.3 (495) | 22.7 (557) | 24.0 (537) |

#### Appendix 4: Unweighted characteristics of the sample and pandemic across pandemic time periods

**Table S2.** Characteristics of the pandemic and the Real-Time Insights COVID-19 in India sample (N=3,662) across the 6 pandemic time periods. Binary and categorical variables are presented using proportions and numbers of respondents; continuous variables are presented using means and interquartile ranges. Cases, deaths, and stringency index shown summarize the mean lagged values (over the three [cases, deaths] months or one [government stringency] month prior to interview date) for all those interviewed during a given pandemic period. Raw cases and deaths are presented per 100,000 population.

| | **Initial lockdown** | **First wave** | **Low cases – Winter 2021** | **Delta wave** | **Low cases – Fall 2021** | **Omicron wave** |
|---|---|---|---|---|---|---|
| N | 4361 | 2896 | 4984 | 2340 | 2444 | 1859 |
| Dates | May 2020 - Jul 2020 | Aug 2020 - Nov 2020 | Dec 2020 - Mar 2021 | Apr 2021 - Jul 2021 | Aug 2021 - Dec 2021 | Dec 2021 - May 2022 |
| Age | 52.1 (35.0 - 68.0) | 52.4 (36.0 - 68.0) | 53.0 (36.0 - 68.0) | 52.9 (36.0 - 68.0) | 53.0 (37.0 - 68.0) | 51.3 (35.0 - 67.0) |
| Female | 47.6 (2078) | 46.4 (1343) | 46.4 (2311) | 46.4 (1085) | 46.6 (1140) | 45.7 (849) |
| Education | | | | | | |
| None | 19.8 (863) | 18.0 (521) | 17.6 (877) | 17.9 (420) | 20.0 (488) | 20.4 (380) |
| Less than secondary | 33.8 (1473) | 35.1 (1016) | 34.4 (1713) | 33.6 (786) | 34.2 (835) | 32.9 (612) |
| Secondary | 28.9 (1261) | 29.5 (854) | 29.9 (1491) | 30.8 (721) | 29.2 (713) | 29.2 (543) |
| Some graduate | 17.5 (764) | 17.4 (505) | 18.1 (903) | 17.6 (413) | 16.7 (408) | 17.4 (324) |
| Rural | 61.4 (2678) | 57.2 (1656) | 57.1 (2847) | 58.3 (1364) | 60.4 (1475) | 62.4 (1160) |
| PHQ score | 0.13 (-0.79 - 0.83) | -0.10 (-0.79 - 0.52) | -0.11 (-0.79 - 0.52) | 0.07 (-0.79 - 0.90) | -0.03 (-0.79 - 0.72) | 0.03 (-0.79 - 0.86) |
| Cases (estimate) | | | | | | |
| Raw | 34.0 (3.3 - 48.2) | 130.2 (86.0 - 170.6) | 68.3 (32.4 - 86.8) | 240.2 (180.7 - 293.5) | 62.8 (5.3 - 81.2) | 623.9 (474.2 - 810.0) |
| Z-score | -0.54 (-0.70 - -0.48) | -0.07 (-0.29 - 0.12) | -0.38 (-0.55 - - 0.29) | 0.47 (0.17 - 0.73) | -0.40 (-0.69 - - 0.31) | 2.35 (1.61 - 3.26) |
| Excess mortality (estimate) | | | | | | |
| Raw | 0.08 (0.00 - 0.11) | 0.45 (0.24 - 0.61) | 0.26 (0.11 - 0.36) | 1.33 (0.53 - 1.89) | 0.48 (0.03 - 0.94) | 0.19 (0.08 - 0.26) |
| Z-score | -0.56 (-0.68 - -0.50) | 0.09 (-0.28 - 0.36) | -0.24 (-0.49 - - 0.08) | 1.58 (0.22 - 2.54) | 0.14 (-0.64 - 0.91) | -0.37 (-0.56 - - 0.24) |
| Cases (admin) | | | | | | |
| Raw | 0.5 (0.0 - 0.4) | 5.6 (1.9 - 7.7) | 5.3 (1.1 - 7.3) | 19.3 (4.0 - 24.8) | 9.8 (0.1 - 4.5) | 11.7 (2.7 - 11.0) |
| Z-score | -0.52 (-0.56 - -0.53) | -0.12 (-0.41 - 0.04) | -0.15 (-0.48 - 0.01) | 0.95 (-0.25 - 1.38) | 0.20 (-0.56 - - 0.21) | 0.35 (-0.35 - 0.30) |
| Deaths (admin) | | | | | | |
| Raw | 0.01 (0.00 - 0.01) | 0.07 (0.02 - 0.11) | 0.05 (0.01 - 0.07) | 0.22 (0.07 - 0.33) | 0.09 (0.00 - 0.11) | 0.10 (0.01 - 0.03) |
| Z-score | -0.45 (-0.54 - -0.48) | -0.04 (-0.39 - 0.26) | -0.19 (-0.46 - - 0.08) | 1.01 (-0.08 - 1.78) | 0.12 (-0.51 - 0.25) | 0.18 (-0.50 - - 0.32) |

| | | | | | | |
|---|---|---|---|---|---|---|
| Stringency index | | | | | | |
| Raw | 91.2 (84.2 - 98.9) | 68.9 (61.5 - 78.9) | 48.3 (40.8 - 56.5) | 66.6 (61.8 - 75.2) | 59.4 (54.6 - 68.2) | 30.5 (20.8 - 40.7) |
| Z-score | 1.28 (0.96 - 1.63) | 0.26 (-0.08 - 0.71) | -0.69 (-1.03 - - 0.31) | 0.15 (-0.07 - 0.55) | -0.18 (-0.40 - 0.22) | -1.50 (-1.95 - - 1.03) |
| Financial impact | | | | | | |
| No impact | 10.1 (440) | 9.2 (266) | 8.8 (437) | 7.4 (174) | 6.4 (157) | 10.7 (199) |
| Too soon to tell | 0.9 (41) | 0.8 (23) | 0.6 (29) | 0.4 (10) | 1.1 (27) | 1.0 (19) |
| Minor impact | 12.7 (556) | 9.1 (263) | 10.9 (545) | 11.2 (261) | 10.1 (248) | 8.8 (164) |
| Moderate impact | 32.7 (1427) | 34.7 (1004) | 34.1 (1699) | 37.6 (880) | 41.0 (1001) | 38.7 (719) |
| Major impact | 43.5 (1897) | 46.3 (1340) | 45.6 (2274) | 43.4 (1015) | 41.4 (1011) | 40.8 (758) |
| Prior household infection | 1.1 (48) | 4.9 (141) | 10.3 (515) | 20.6 (483) | 25.6 (625) | 29.1 (541) |

### Appendix 5: Estimated coefficients for pandemic markers, including coefficients from a model adjusted for all other pandemic markers

**Table S3.** Estimated coefficients for the association between pandemic markers and PHQ-4 score in the Real-Time Insights of COVID-19 in India sample (N=3,662) for crude models, models adjusted for demographics (age [spline], gender, educational attainment, and rural/urban residence), and models adjusted for demographics and all other pandemic markers. All coefficients are shown as mean differences.

| | **Crude** | **Adjusted for demographics** | **Full adjustment** |
|---|---|---|---|
| Cases - IHME | 0.019 (0.007 - 0.031)** | 0.019 (0.007 - 0.031)** | 0.042 (0.029 - 0.056)*** |
| Deaths - IHME | 0.041 (0.030 - 0.053)*** | 0.041 (0.030 - 0.053)*** | 0.016 (0.004 - 0.029)** |
| Stringency index | 0.060 (0.048 - 0.072)*** | 0.060 (0.048 - 0.072)*** | 0.083 (0.070 - 0.096)*** |
| Financial impact | | | |
| Too soon to tell | 0.195 (0.047 - 0.343)** | 0.195 (0.047 - 0.343)** | 0.200 (0.051 - 0.350)** |
| Minor impact | 0.062 (0.017 - 0.108)** | 0.062 (0.017 - 0.108)** | 0.070 (0.025 - 0.116)** |
| Moderate impact | 0.241 (0.201 - 0.282)*** | 0.241 (0.201 - 0.282)*** | 0.252 (0.211 - 0.292)*** |
| Major impact | 0.447 (0.406 - 0.489)*** | 0.447 (0.406 - 0.489)*** | 0.454 (0.412 - 0.496)*** |
| Prior household infection | 0.113 (0.068 - 0.158)*** | 0.113 (0.068 - 0.158)*** | 0.134 (0.090 - 0.178)*** |

*p<0.05, **p<0.01, ***p<0.001

#### Appendix 6: Statistical significance of effect modification by age group, gender, and rural/urban residence

**Table S4.** Statistical significance of effect modification of the association between pandemic characteristics and PHQ4 score by age group, gender, and effect modification. Statistical significance was tested using chi-squared tests comparing the model with the interaction terms to a model without the interaction terms. All models controlled for demographic characteristics (age [spline], gender, educational attainment, and rural/urban residence).

| | P-value |
|---|---|
| **Age category** | |
| Time period | <0.001 |
| Cases - IHME | <0.001 |
| Deaths - IHME | <0.001 |
| Stringency index | <0.001 |
| Cases - Admin | <0.001 |
| Deaths - Admin | <0.001 |
| Financial impact | <0.001 |
| Prior household infection | <0.001 |
| **Gender** | |
| Time period | 0.003 |
| Cases - IHME | <0.001 |
| Deaths - IHME | 0.123 |
| Stringency index | 0.038 |
| Cases - Admin | 0.006 |
| Deaths - Admin | 0.169 |
| Financial impact | <0.001 |
| Prior household infection | 0.017 |
| **Rural/urban** | |
| Time period | 0.037 |
| Cases - IHME | 0.476 |
| Deaths - IHME | <0.001 |
| Stringency index | 0.635 |
| Cases - Admin | 0.301 |
| Deaths - Admin | 0.902 |
| Financial impact | 0.545 |
| Prior household infection | 0.548 |

#### Appendix 7: Sensitivity analyses using binary outcome instead of continuous score

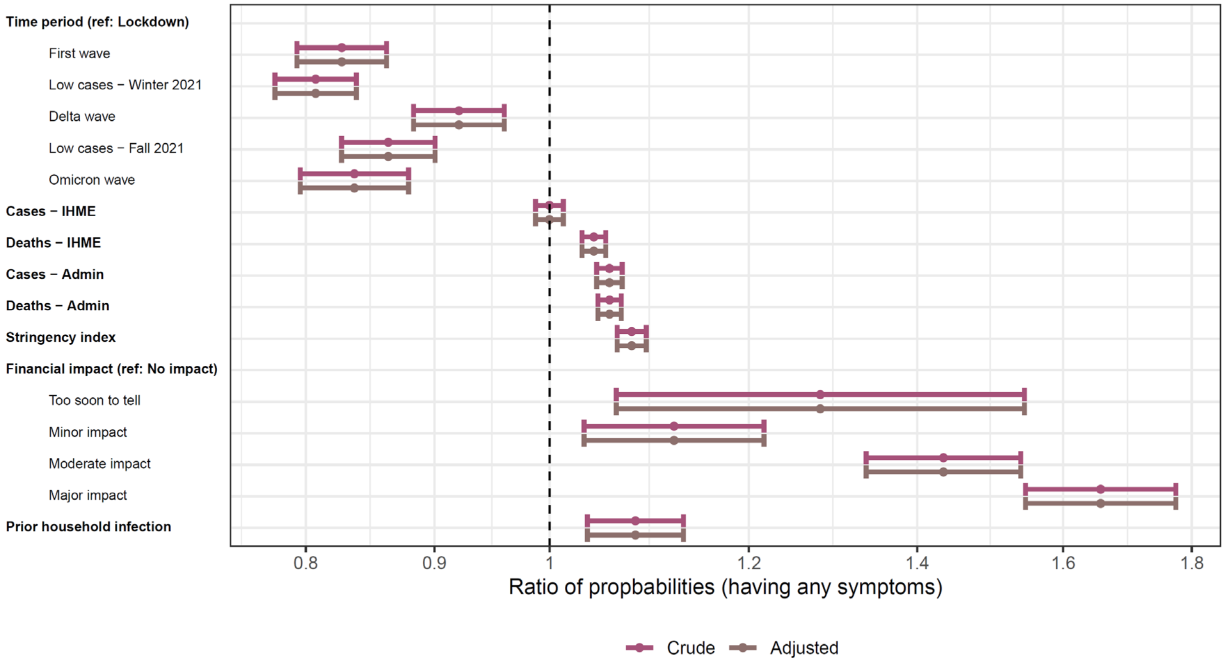

**Figure S2.** Associations between pandemic periods and pandemic markers and reporting of any PHQ4 symptom in the Real-Time Insights COVID-19 in India sample (N=3,662). Associations with all continuous markers (cases, deaths, government stringency index) are shown as associations for 1 SD unit difference. Adjusted models are adjusted for age (spline), gender, educational attainment, and rural/urban residence.

**Table S5.** Estimated coefficients for the association between pandemic markers and reporting of any PHQ4 symptom in the Real-Time Insights COVID-19 in India sample (N=3,662) for crude models, models adjusted for demographics (age [spline], gender, educational attainment, and rural/urban residence), and models adjusted for demographics and all other pandemic markers. All coefficients are shown as prevalence ratios.

| | **Crude** | **Adjusted for demographics** | **Full adjustment** |
|---|---|---|---|
| Cases - IHME | 1.000 (0.987 - 1.013) | 1.000 (0.987 - 1.013) | 1.026 (1.010 - 1.041)*** |
| Deaths - IHME | 1.041 (1.030 - 1.053)*** | 1.041 (1.030 - 1.053)*** | 1.021 (1.009 - 1.033)*** |
| Stringency index | 1.078 (1.064 - 1.093)*** | 1.078 (1.064 - 1.093)*** | 1.095 (1.079 - 1.112)*** |
| Financial impact | | | |
| Too soon to tell | 1.281 (1.063 - 1.544)** | 1.281 (1.063 - 1.544)** | 1.299 (1.076 - 1.568)** |
| Minor impact | 1.121 (1.032 - 1.217)** | 1.121 (1.032 - 1.217)** | 1.129 (1.040 - 1.226)** |
| Moderate impact | 1.434 (1.336 - 1.539)*** | 1.434 (1.336 - 1.539)*** | 1.458 (1.359 - 1.563)*** |
| Major impact | 1.656 (1.546 - 1.774)*** | 1.656 (1.546 - 1.774)*** | 1.673 (1.562 - 1.792)*** |
| Prior household infection | 1.082 (1.035 - 1.130)*** | 1.082 (1.035 - 1.130)*** | 1.116 (1.068 - 1.165)*** |

*p<0.05, **p<0.01, ***p<0.001

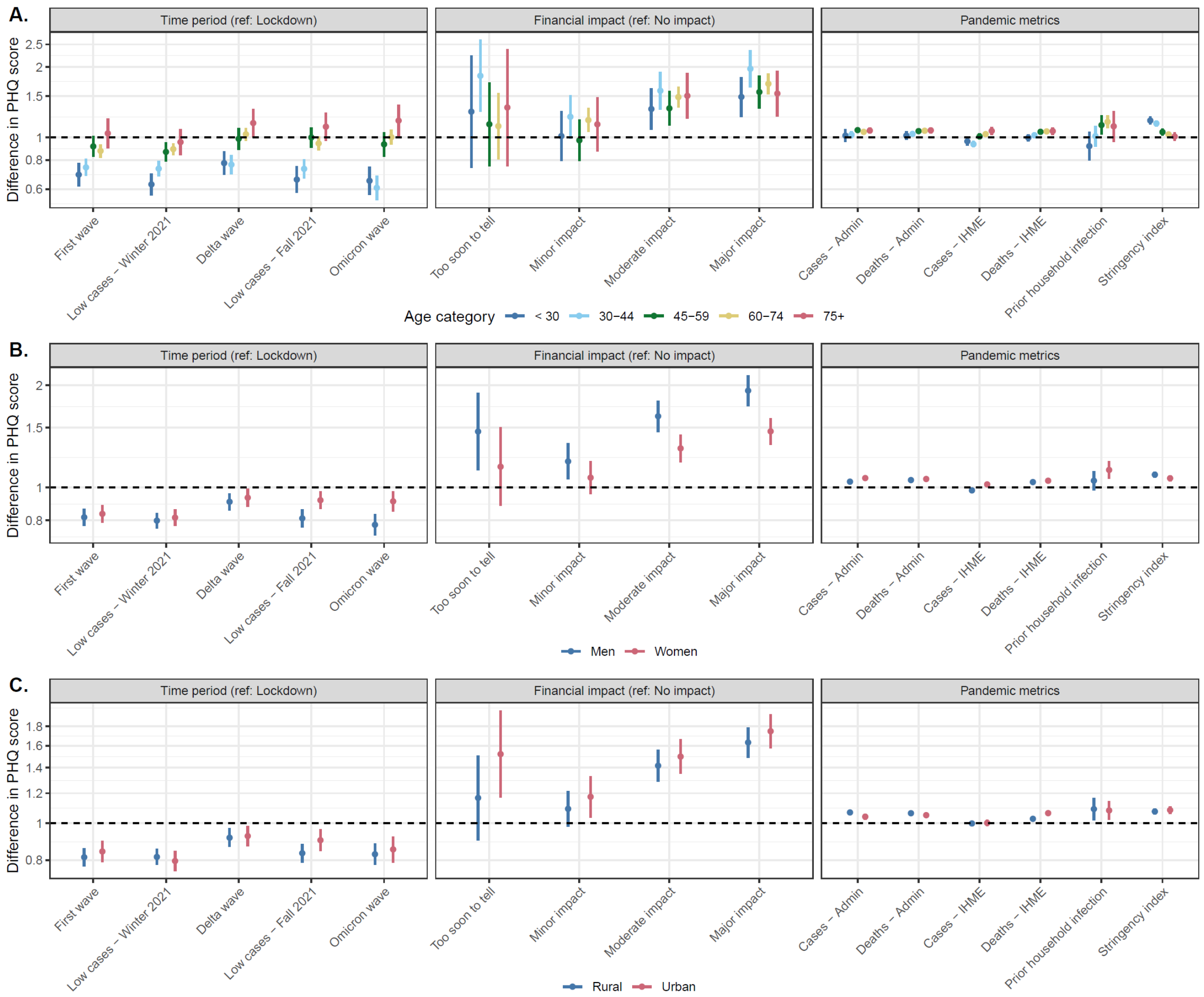

**Figure S3.** Effect modification of the association between COVID-19 pandemic periods and COVID-19 pandemic markers and reporting of any PHQ4 symptom by age group [A], gender [B], and urban/rural residence [C] in the Real-Time Insights COVID-19 in India sample (N=3,662). All models are adjusted for age, gender, educational attainment, and rural/urban residence.

### Appendix 8: Sensitivity analyses using weighted regression models

Results from models using analytic weights were shifted closer to findings observed in younger adults because the RTI-COVID India sample contains a larger proportion of older adults compared to the general population of India due to the sampling frame and targeted recruitment of LASI-DAD participants. This leads to changes in the statistical significance of individual effects for administrative cases, self-reported minor financial impact and prior household infection, although the direction of estimated coefficients remained the same. Additionally, the lockdown period stands out further in this analysis as being associated with higher mental health symptom burden compared to subsequent time periods. However, effect modification results by age are consistent, further illustrating that observed differences between younger and older adults are driving differences between unweighted and weighted estimates. Additionally, other than changes in statistical significance for self-reported minor-financial impact and estimated deaths, there were no large differences in associations for mutually-adjusted pandemic characteristics.

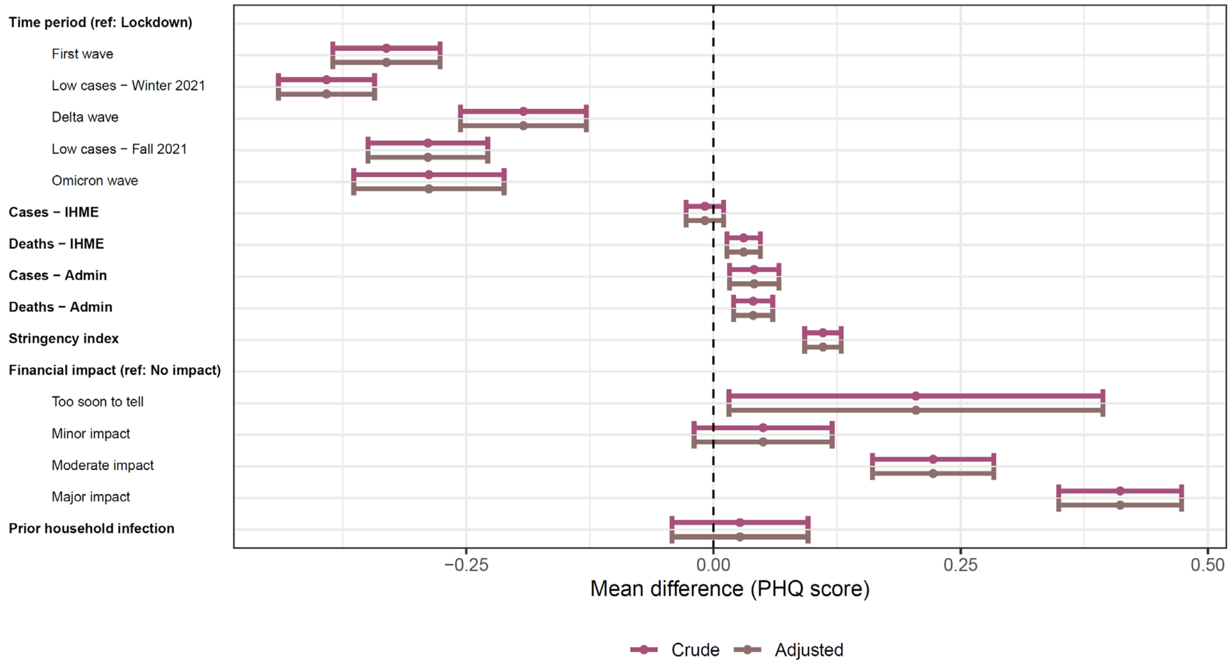

**Figure S4.** Weighted associations between COVID-19 pandemic periods and COVID-19 pandemic markers and PHQ-4 scores in the Real-Time Insights COVID-19 in India sample (N=3,662). Associations with all continuous markers (cases, deaths, government stringency index) are shown as associations for 1 SD unit difference. Adjusted models are adjusted for age (spline), gender, educational attainment, and rural/urban residence.

**Table S6.** Estimated coefficients for the association between pandemic markers and PHQ-4 score in the Real-Time Insights COVID-19 in India sample (N=3,662) for weighted models (crude, adjusted for demographics [age [spline], gender, educational attainment, and rural/urban residence], and adjusted for demographics and all other pandemic markers). All coefficients are shown as mean differences.

| | **Crude** | **Adjusted for demographics** | **Full adjustment** |
|---|---|---|---|
| Cases - IHME | -0.009 (-0.028 - 0.010) | -0.009 (-0.028 - 0.010) | 0.048 (0.027 - 0.069)*** |
| Deaths - IHME | 0.031 (0.014 - 0.047)*** | 0.031 (0.014 - 0.047)*** | 0.010 (-0.008 - 0.028) |
| Stringency index | 0.111 (0.092 - 0.129)*** | 0.111 (0.092 - 0.129)*** | 0.132 (0.113 - 0.152)*** |
| Financial impact | | | |
| Too soon to tell | 0.205 (0.016 - 0.394)* | 0.205 (0.016 - 0.394)* | 0.218 (0.027 - 0.410)* |
| Minor impact | 0.050 (-0.020 - 0.120) | 0.050 (-0.020 - 0.120) | 0.055 (-0.014 - 0.124) |
| Moderate impact | 0.222 (0.161 - 0.284)*** | 0.222 (0.161 - 0.284)*** | 0.233 (0.172 - 0.295)*** |
| Major impact | 0.411 (0.349 - 0.473)*** | 0.411 (0.349 - 0.473)*** | 0.410 (0.348 - 0.472)*** |
| Prior household infection | 0.027 (-0.042 - 0.096) | 0.027 (-0.042 - 0.096) | 0.094 (0.026 - 0.163)** |

*p<0.05, **p<0.01, ***p<0.001

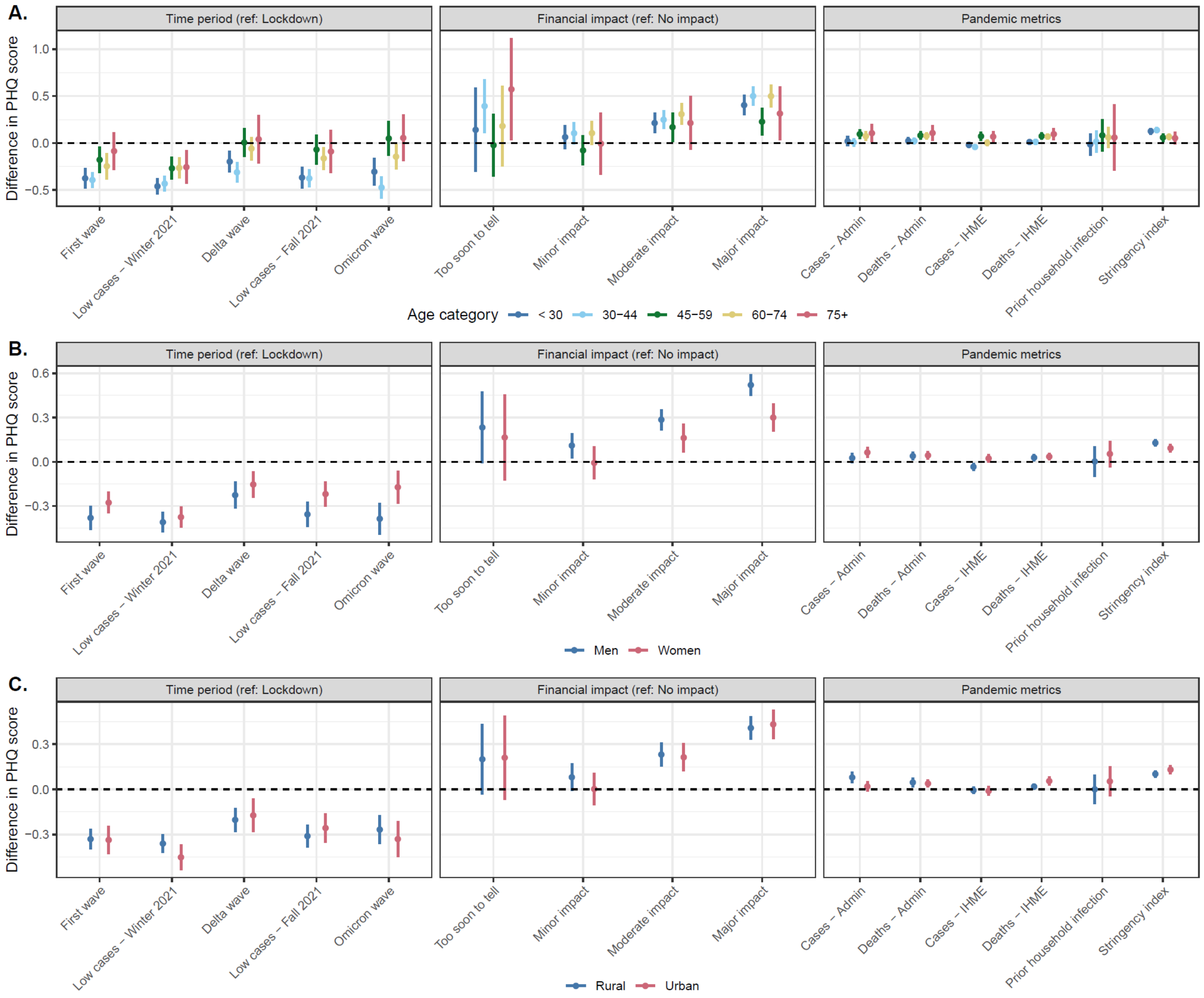

**Figure S5.** Effect modification of the association between COVID-19 pandemic periods and COVID-19 pandemic markers and PHQ4 score by age group [A], gender [B], and urban/rural residence [C] in the Real-Time Insights COVID-19 in India sample (N=3,662). All models are weighted and adjusted for age, gender, educational attainment, and rural/urban residence.
